# Effects of Starting and Stopping Combined Oral Contraceptives on Markers of Ovarian Reserve

**DOI:** 10.64898/2026.05.29.26354411

**Authors:** U Bernig, M Kördel, I Sundström-Poromaa, N.B. Kroemer, M Henes

## Abstract

**Objective:** To examine the effects of combined oral contraceptive (OC) use on clinical markers of ovarian reserve by comparing Anti-Muellerian Hormone (AMH), antral follicle count (AFC), and ovarian volume (OV) before and after starting or stopping OC.

**Methods:** This analysis is based on data from a prospective cohort study conducted at the University Hospital Tübingen, Germany, as part of the IRTG-2804 project. A total of 54 healthy women were included and categorized into three groups based on their OC use status: OC starters (*n* = 12), stoppers (*n* = 16), and long-term OC-users (*n* = 26). Each participant underwent a transvaginal ultrasound (including AFC and OV) and serum sampling (including AMH) at two time points (S1 and S2), three to six months apart. OC starters were assessed first during the early follicular phase (day 1–7) and then during active OC intake (day 8–21), while stoppers were assessed in the reverse order. Long-term users were assessed twice during active OC intake.

**Results:** OC stoppers showed significant within-group increases in all ovarian reserve markers, including AMH (Δ = 2.57 ng/mL, *p* < .001), AFC (Δ = 3.88, *p* = .004), and OV, which almost doubled (1.94-fold increase; 95% CI [1.35, 2.80], *p* < .001). In contrast, OC starters exhibited a significant decline in AMH (Δ = −1.25 ng/mL, *p* = .013), but no changes in AFC or OV. No significant longitudinal changes were observed among long-term OC users.

**Conclusion:** AMH levels decrease after starting OC use whereas AFC and OV are not affected. In contrast, AMH, AFC, and OV recover within three to six months after stopping OC, suggesting a reversible suppression of ovarian reserve markers during OC use. These findings are clinically relevant for fertility counselling and for the interpretation of ovarian reserve markers in women using hormonal contraception.

## Introduction

Ovarian reserve reflects the remaining pool of follicles in the ovaries and represents a key determinant of female reproductive potential (Gleicher et al., 2011; Tal & Seifer, 2017). As the true ovarian reserve, meaning the pool of non-growing follicles, cannot be directly assessed in clinical practice, surrogate markers such as anti-Müllerian hormone (AMH), antral follicle count (AFC), and ovarian volume (OV) are routinely used to estimate functional ovarian reserve (Hansen et al., 2011; La Marca et al., 2010). Among these, AMH and AFC are considered the most reliable indicators (Penzias et al., 2020). Ovarian reserve assessment is most commonly used in the context of assisted reproductive technologies (ART), such as in vitro fertilization (IVF) or intracytoplasmic sperm injection (ICSI), where these markers serve to estimate the expected ovarian response to controlled ovarian stimulation and to guide treatment planning. Since these markers directly influence clinical decision-making, it is crucial to understand factors that may alter their levels.

While age is the primary determinant of ovarian reserve, several modifiable and non-modifiable factors have been associated with reduced AMH levels, including autoimmune diseases, ovarian surgery, endometriosis, smoking, chemotherapy, vitamin D deficiency and genetic variants (Tal & Seifer, 2017). For example, women with rheumatic diseases have been reported to show significantly lower AMH concentrations than healthy controls (Henes et al., 2015; Lawrenz et al., 2011; Pecher et al., 2024), and surgical removal of ovarian cysts, particularly endometriomas, has been linked to a postoperative decline in AMH (Alammari et al., 2017; Henes et al., 2018). In contrast, women with polycystic ovary syndrome (PCOS) often show elevated AMH levels and an increased AFC (Dewailly et al., 2011; Pigny et al., 2006). These observations underline that ovarian reserve markers may be affected by factors other than ovarian aging itself. Among these potential modifiers, exogenous hormonal exposure, particularly oral contraceptive (OC) use, has attracted increasing attention since the late 2000s.

OC are among the most commonly used contraceptive methods in women of reproductive age in Germany (Scharmanski, S. & Hessling, A., 2024). Most contemporary OC contain ethinyl estradiol in combination with a synthetic progestin, administered in either monophasic or multiphasic regimens. By suppressing the hypothalamic-pituitary-ovarian axis through negative feedback mechanisms, OC inhibit gonadotropin secretion, follicular recruitment, and ovulation (Genazzani et al., 2023; Rivera et al., 1999). Their widespread use and potent endocrine effects have raised questions regarding their influence on ovarian physiology beyond contraception. Indeed, several cross-sectional studies have demonstrated significantly lower AMH levels, AFC, and OV in OC users compared with naturally cycling women (Bentzen et al., 2012; Birch Petersen et al., 2015; Landersoe, Forman, et al., 2020; Nelson, Ewing, et al., 2023). Longitudinal studies suggest that these parameters begin to recover within a few months after stopping OC (Landersoe, Birch Petersen, et al., 2020; Letourneau et al., 2017; van den Berg et al., 2010). However, studies investigating the impact of starting OC on ovarian reserve markers have yielded inconsistent results (Andersen et al., 2011; Arbo et al., 2007; Kallio et al., 2013; Li et al., 2011; Somunkiran et al., 2007). Thus, while OC use appears to influence clinical markers of ovarian reserve, the magnitude and direction of these changes across different OC use statuses (start, stop, and continued use) remain incompletely understood.

Most available larger studies have compared long-term users with non-users (Bentzen et al., 2012; Bernardi et al., 2021; Birch Petersen et al., 2015; Landersoe, Forman, et al., 2020; Nelson, Ewing, et al., 2023). To our knowledge, no previous study has directly compared women starting and stopping OC within the same longitudinal study framework. Moreover, the available longitudinal evidence on changes in ovarian reserve markers after starting OC remains limited by small sample sizes, short follow-up intervals, and methodological heterogeneity (Arbo et al., 2007; Kallio et al., 2013; Li et al., 2011; Somunkiran et al., 2007; Streuli et al., 2008). Consequently, it remains unclear whether lower levels of ovarian reserve markers observed during OC use mainly represent a transient, reversible suppression or more sustained alterations in ovarian physiology.

To address this gap, we conducted a prospective cohort study investigating mid-term (3–6 months) changes in AMH, AFC, and OV in women starting, stopping, or continuing OC use. By longitudinally assessing these markers over a defined follow-up period, the study aims to clarify the magnitude, trajectory, and reversibility of OC-associated changes in clinical markers of ovarian reserve.

## Methods

### Study design and study population

The present analysis is based on data from a larger study examining the association between hormonal contraception and stress reactivity, which began data collection in February 2024. Premenopausal women aged 18–40 years with a BMI between 18 and 35 kg/m² were eligible for inclusion if they were planning to start, stop or continue the use of a monophasic OC. The choice of a hormone-free interval (HFI) with withdrawal bleeding during the pill cycle was optional. Participants in the stopper and long-term user groups had to use OC for at least six months prior to study enrolment. Women intending to start OC use were required to have a regular natural menstrual cycle (25–35 days). Exclusion criteria included psychiatric, neurological, endocrine, or severe physical illness, current or recent pregnancy or breastfeeding (within the past 12 months), regular medication use (except OC), use of hormonal contraceptives other than OCs (e.g., progestin-only pills, hormonal intrauterine devices, vaginal rings) or copper intrauterine devices and regular smoking.

Participants were assessed at two time points (S1 and S2) according to their OC use status (Figure 1). Participants were classified as starters (women who started OC use), stoppers (women who stopped OC use), and long-term users (women who continued OC use).

**Figure 1.**
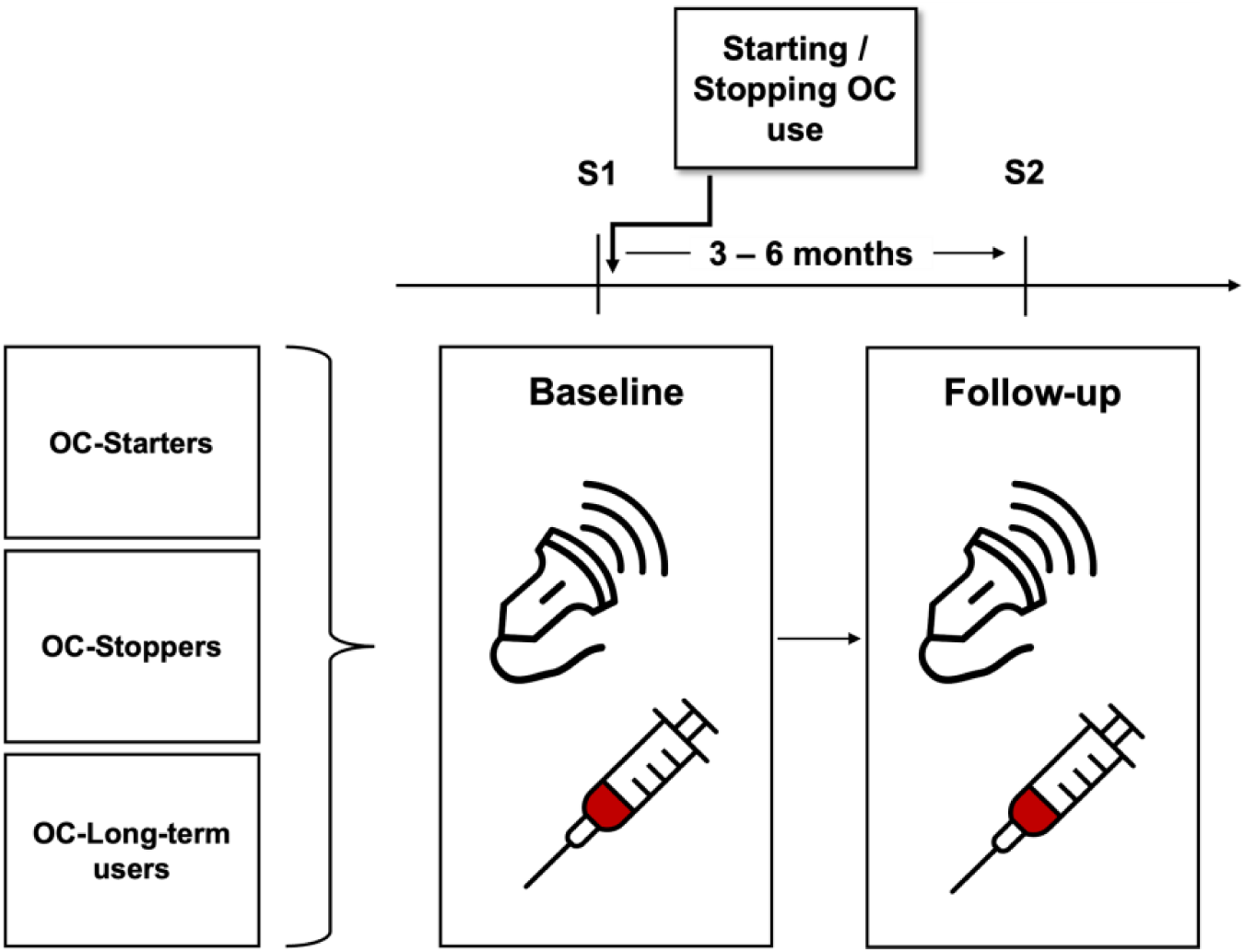
Study design. *Note.* Participants were classified as starters, stoppers, or long-term users of OC. Assessments were performed at baseline (S1) and follow-up (S2), separated by approximately 3–6 months. Ultrasound examinations and blood sampling were conducted at both time points.

For starters, S1 assessments were conducted during the early follicular phase (day 1–7 of the menstrual cycle), whereas S2 assessments took place during active OC use (day 8–21). For stoppers, S1 assessments were performed during active OC use, and S2 assessments during the early follicular phase after at least three spontaneous menstrual cycles. For long-term users, both S1 and S2 assessments were conducted during active OC use. Depending on cycle recovery and scheduling availability, the interval between assessments ranged from approximately three to six months.

At S1, participants completed a questionnaire regarding past, current, or planned OC use. Data collected included OC formulation, duration of use, age at first prescription, regimen (with/without HFI), reason for starting and if applicable reason for stopping OC use.

### Assessment of ovarian reserve parameters

At both time points, a standardized transvaginal B-mode ultrasound (Voluson E, GE Healthcare, 4–10 MHz transducer) was performed at the Department of Gynecology, University Hospital Tübingen. OV and AFC were assessed according to a predefined study protocol. AFC was defined as the total number of follicles measuring 2–10 mm across both ovaries, in accordance with Broekmans et al. (2010). OV for each ovary was calculated using the formula for a prolate ellipsoid: *V = d₁ × d₂ × d₃ × π/6* (Cohen et al., 1990; Rosendahl et al., 2010). The mean OV was defined as the average of both ovaries (Landersoe, Birch Petersen, et al., 2020; Rosendahl et al., 2010). All examinations were conducted by UB in collaboration with experienced physicians from the Division of Endocrinology and Reproductive Medicine at the University Hospital Tübingen. Structural abnormalities were documented and, if necessary, reviewed by the study’s principal investigator.

Additionally, venous blood samples (serum) were collected at S1 and S2. AMH levels were quantified at the central laboratory of the University Hospital Tübingen using an electrochemiluminescence immunoassay (Elecsys® AMH Plus Assay, Roche). The validated measuring range of the assay is 0.01–23 ng/mL (Roche Diagnostics, 2024). Age-specific reference intervals for women aged 20–39 years range from 0.405 to 9.95 ng/mL, corresponding to the 5th to 95th percentiles reported by the manufacturer (Roche Diagnostics, 2024).

### Statistical analysis and software

All statistical analyses were conducted using *Jamovi* version 2.6 (The jamovi project, 2025) and *R* versions 4.3.2 (R Core Team, 2023) and 4.4.3 (R Core Team, 2025). Descriptive statistics were performed in Jamovi, while linear mixed-effects models (LMMs) were conducted in R. A significance level of *p* < .05 was applied. No matching was performed, as potential confounding was addressed by including covariates in the statistical models. Age was included as an a priori covariate given its well-established association with ovarian reserve parameters, whereas BMI was included to account for its potential influence on these measures. Continuous covariates were grand-mean centered based on baseline (S1) values. OV was log-transformed prior to analysis to approximate normality.

LMMs with random intercepts for participants (ID) were used to assess group-by-time interactions. Fixed effects included time (S1 vs. S2), group (reference: long-term OC users), their interaction and the covariates age and BMI. All available observations were included, and missing data were not imputed. Models were estimated using restricted maximum likelihood (REML). *t*-tests and *p*-values were derived using Satterthwaite approximations, and confidence intervals were based on Wald tests.

To facilitate interpretation, estimated marginal means (EMMs) adjusted for centered age and BMI were derived from the LMMs. Estimates reflect contrasts at the average age and BMI of the total sample at S1. Within-group contrasts (S2 vs. S1) indicate whether AMH, AFC or OV changed significantly over time, while between-group differences are instead captured by the interaction effects reported in Table S3 in the supplementary material.

### Ethics statement

The study was approved by the local ethics committee of the Eberhard-Karls-University Tübingen (IRB-equivalent; approval number: 412/2023BO1) and all participants gave their written informed consent before the experiment. They received monetary compensation. The study was preregistered at www.clinicaltrials.gov (trial no. NCT06223126). All procedures were carried out in accordance with the Declaration of Helsinki.

## Results

### Sample characteristics

For the current analyses, the sample included 54 participants with 12 starters, 16 stoppers and 26 long-term users. Figure 2 illustrates the distribution of participants across study visits and their final inclusion in the analyses.

**Figure 2.**
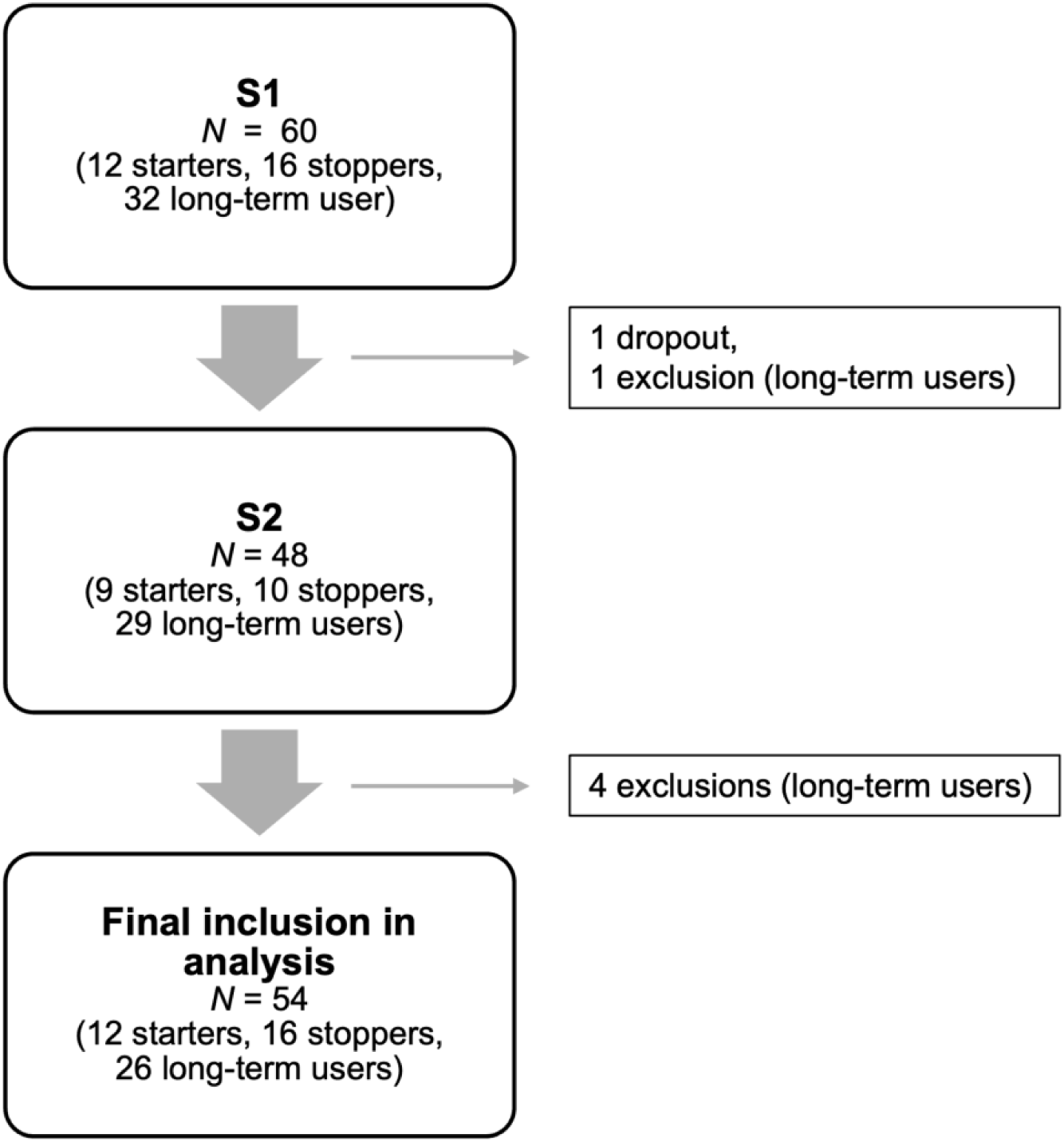
Flowchart of the study sample across assessment time points S1 and S2 until final inclusion in the analysis. *Note.* At the time of analysis, not all participants had completed both sessions (with some not yet assessed at S1 and others not yet at S2), which explains the discrepancies in sample sizes across time points. Dropouts, participants not meeting the study criteria, and long-term users with incomplete datasets were excluded from analysis, while starters and stoppers with valid S1 data but pending S2 assessments were retained in order to maintain an adequate sample size.

Participants were aged 19 to 36 years (*M* = 24.7 years, *SD* = 3.9) with a mean BMI of 22.5 kg/m² (*SD* = 2.3). Most participants reported German as their native language (*n* = 50), all had completed secondary education qualifying for university admission (*n* = 54) and none had children. No participant reported regular medication intake relevant to ovarian reserve markers other than OC use. Reported gynecological conditions included dysmenorrhea (*n* = 17), acne (*n* = 5) and endometriosis with heavy menstrual bleeding (*n* = 1). One stopper reported rheumatoid arthritis, not requiring medication. Median OC duration at S1 was 7.5 years (range = 3.6–16.0) in stoppers and 5.9 years (range = 0.8–11.6) in long-term users. Most participants in all groups followed a 21/7 regimen. Baseline characteristics and information on OC use are shown in Table 1.

**Table 1.** Baseline Information and Characteristics of OC Use.

|  | Starters | Stoppers | Long-term users |
| --- | --- | --- | --- |
| **Age (years)** |  |  |  |
| $M \pm SD$ | $24.3 \pm 3.9$ | $24.9 \pm 3.4$ | $24.8 \pm 4.2$ |
| Min–Max | 19–32 | 19–30 | 20–36 |
| **BMI ($\text{kg/m}^2$)** |  |  |  |
| $M \pm SD$ | $21.6 \pm 1.9$ | $23.0 \pm 2.6$ | $22.6 \pm 2.3$ |
| Min–Max | 18.9–24.6 | 19.8–27.5 | 19.1–27.4 |
| **Age at menarche (years)** |  |  |  |
| $M \pm SD$ | $13.1 \pm 0.9$ | $12.9 \pm 1.7$ | $13.2 \pm 1.1$ |
| Min–Max | 12–15 | 9–15 | 11–15 |
| **Age at first OC-prescription (years)** |  |  |  |
| $M \pm SD$ | $20.2 \pm 5.3$ | $15.9 \pm 1.3$ | $17.0 \pm 1.9$ |
| **OC-regimen** |  |  |  |
| $n$ (%) | | | |
| Continuous | 2 (17) | 3 (19) | 4 (15) |
| HFI („pill break“) | 10 (83) | 13 (81) | 22 (85) |
| **EE dose in OC ($\mu\text{g}$)** |  |  |  |
| $n$ (%) | | | |
| 20 | 4 (33) | 2 (12) | 10 (38) |
| 30 | 7 (58) | 14 (88) | 16 (62) |
| 35 | 1 (8) | 0 (0) | 0 (0) |
| **Progestin generation $n$ (%)** |  |  |  |
| 2nd generation | 7 (58) | 6 (37) | 12 (46) |
| 3rd generation | 0 (0) | 0(0) | 1 (4) |
| 4th generation | 5 (42) | 10 (63) | 13 (50) |

*Note.* EE = Ethinylestradiol. HFI = hormone-free interval. % = percentage relative to the respective group. Characteristics of OC use for the group of OC starters refer to their planned intake, as they had not yet started OC use at S1.

### Descriptive patterns of ovarian reserve markers by study group and timepoint

Descriptive statistics for ovarian reserve markers are presented first to provide an overview of group-specific patterns across the two study timepoints. Table 2 displays descriptive values for all three groups at both measurement timepoints (S1 and S2, ∼3–6 months apart), including AMH, AFC and mean OV. Descriptive values for right and left OV are provided in Supplementary Table S1. Overall, starters showed declines across all markers. In contrast, stoppers showed higher median values at follow-up, with median AMH nearly doubling from 2.48 to 4.79 ng/mL and median OV rising from 2.6 to 5.2 mL. In long-term users, median values were slightly lower at S2 than at S1 across all parameters. Since not all measurements were available for every participant, sample sizes vary across parameters as indicated in the table notes. Exact sample sizes for each marker, group, and assessment are provided in Supplementary Table S2.

**Table 2.** Ovarian reserve markers before and 3 to 6 months after OC start, stop, or continuous use.

|  | Starters |  | Stoppers |  | Long-term users |  |
| --- | --- | --- | --- | --- | --- | --- |
|  | S1 | S2 | S1 | S2 | S1 | S2 |
| **Participants *n*** | 12 | 9 | 16 | 10 | 26 | 26 |
| **AMH (ng/mL)** |  |  |  |  |  |  |
| *Md* | 3.29 | 2.57 | 2.48 | 4.79 | 2.83 | 2.46 |
| Min–Max | 1.29–9.10 | 1.26–3.78 | 0.66–7.50 | 2.22–12.10 | 0.55–8.17 | 0.68–7.26 |
| **AFC (n)** |  |  |  |  |  |  |
| *Md* | 11 | 9 | 11 | 12.5 | 9.5 | 8 |
| Min–Max | 7–15 | 6–13 | 4–16 | 9–20 | 5–19 | 5–20 |
| **Mean OV (mL)** |  |  |  |  |  |  |
| *Md* | 4.8 | 3.3 | 2.6 | 5.2 | 2.9 | 2.0 |
| Min–Max | 2.7–9.6 | 2.6–9.0 | 0.9–5.4 | 2.2–9.1 | 1.0–6.5 | 1.2–8.1 |

*Note.* For ultrasound-based outcomes, sample sizes varied by marker and assessment. Ultrasound examination was not feasible in two starters at S1, one stopper at S1, and one stopper at S2. In addition, AFC could not be assessed in one stopper at either assessment. Ovarian volume values at S2 were excluded in two participants because of ovarian cysts.

### Between-group longitudinal analyses show increases in ovarian reserve markers among OC stoppers

To examine whether longitudinal changes in ovarian reserve markers differed between study groups, linear mixed models were fitted for AMH, AFC, and mean OV, with time, group, and their interaction as predictors and with centered age and centered BMI included as covariates. A significant time-by-group interaction was observed for AMH, indicating a greater increase in stoppers (*b* = 2.85, *t* = 5.34, *p* < .001) relative to long-term users, while starters showed a non-significant decrease (*b* = −0.97, *t* = −1.73, *p* = .091). For AFC, stoppers also demonstrated a significantly greater increase relative to long-term users, corresponding to approximately five follicles (*b* = 5.38, *t* = 3.65, *p* < .001), while long-term users showed a decrease that narrowly missed statistical significance (*b* = −1.50, *t* = −2.00, *p* = .054). Similar patterns emerged for log-transformed OV, with a significant increase in stoppers (*b* = 0.85, *t* = 4.09, *p* < .001) and no significant changes in starters. Detailed estimates are provided in the supplementary material (Table S3). Marginal *R²* values were .14 (AMH), .20 (AFC), and .23 (OV), with a conditional *R²* of .79, .49, and .60, respectively. Residual diagnostics supported normality assumptions for all models after log transformation for OV.

### Within-group longitudinal changes in ovarian reserve markers show increases in stoppers and decreases in starters

To further characterize the direction and magnitude of change within each study group, EMMs were used to derive within-group contrasts from S1 to S2 for AMH, AFC, and mean OV (Figure 3). AMH significantly increased in stoppers (Δ = 2.57 ng/mL, *SE* = 0.45, 95% CI [1.66, 3.49], *t*(45.7) = 5.68, *p* < .001) and decreased in starters (Δ = −1.25 ng/mL, *SE* = 0.48, 95% CI [−2.22, −0.28], *t*(44.4) = −2.59, *p* = .013), while no significant change was observed in long-term users (Δ = −0.28 ng/mL, *SE* = 0.29, 95% CI [−0.86, 0.30], *t*(42.1) = −0.98, *p* = .333).

**Figure 3.**
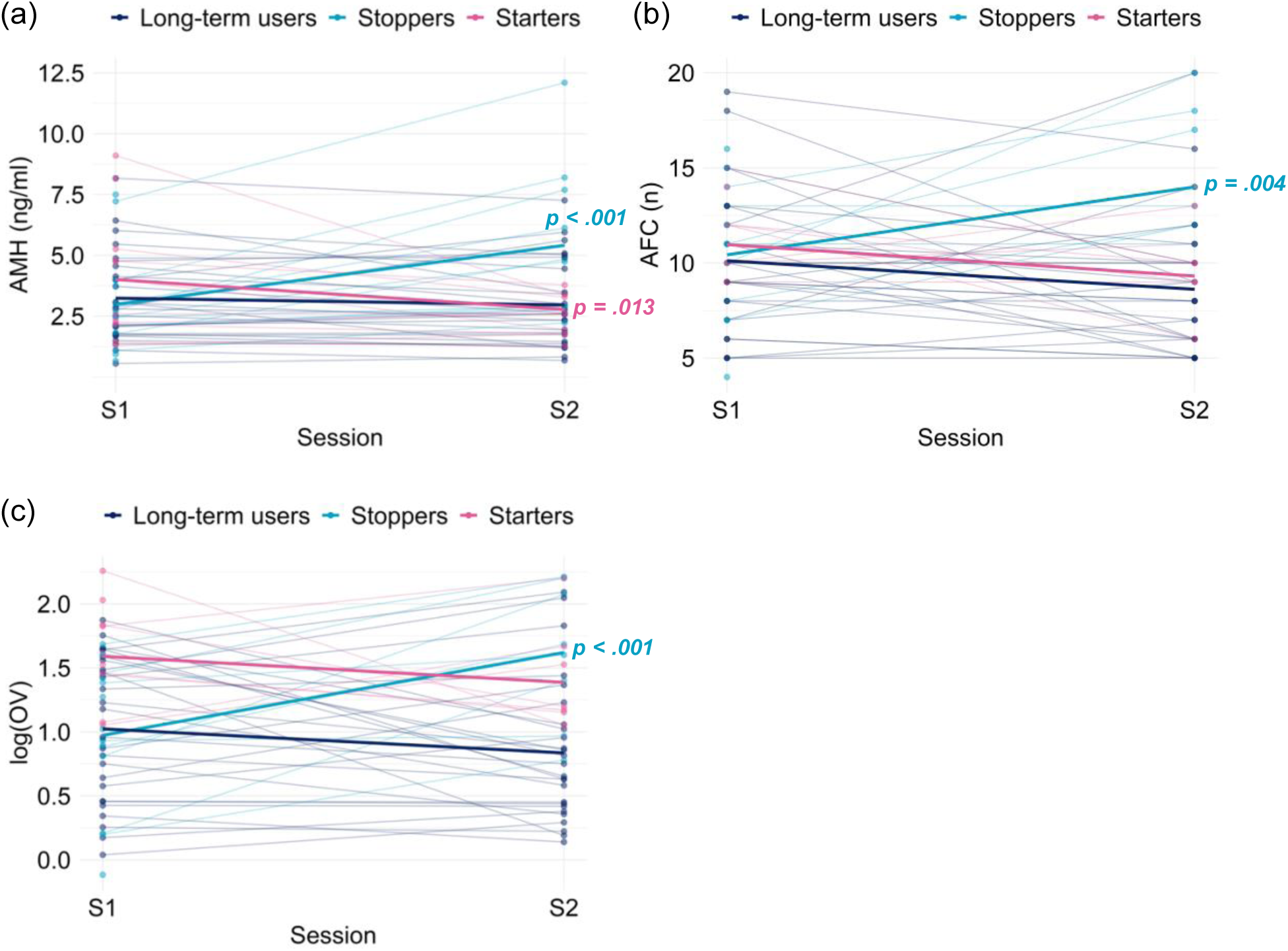
Change in ovarian reserve markers between sessions S1 and S2 stratified by group. *Note.* (a) AMH, (b) AFC, (c) Ovarian volume (OV). Individual trajectories are shown as thin lines, and group-specific means estimated by linear mixed models (EMMs adjusted for age and BMI) as thick lines. OV values were log-transformed (raw values in mL). Among stoppers, significant increases were observed in model-based mean AMH (*p* < .001), AFC (*p* = .004), and OV (*p* < .001) from S1 to S2, whereas starters showed a significant decrease in AMH (*p* = .013).

AFC significantly increased in stoppers (Δ = 3.88, *SE* = 1.28, 95% CI [1.31, 6.44], *t*(49.3) = 3.04, *p* = .004). No significant changes were observed in long-term users (Δ = −1.50, *SE* = 0.75, 95% CI [−3.02, 0.02], *t*(39.8) = −1.99, *p* = .053) or starters (Δ = −1.71, *SE* = 1.29, 95% CI [−4.32, 0.89], *t*(46.0) = −1.32, *p* = .192).

For OV, stoppers showed a significant rise in log-transformed values (Δ = 0.67, *SE* = 0.18, 95% CI [0.30, 1.03], *t*(47.7) = 3.67, *p* < .001), corresponding to a 1.94-fold increase after back-transformation (95% CI [1.35, 2.80]). No significant changes were observed in long-term users (Δ = −0.19, *SE* = 0.11, 95% CI [−0.40, 0.03], *t*(38.6) = −1.77, *p* = .085) or starters (Δ = −0.20, *SE* = 0.19, 95% CI [−0.59, 0.19], *t*(46.5) = −1.02, *p* = .312).

## Discussion

Previous research has shown that OC use is associated with alterations in markers of ovarian reserve. However, it remains unclear to what extent these changes differ depending on starting, stopping, or long-term OC use, and how these markers reflect dynamic ovarian suppression and recovery over time.

By using a longitudinal study design and directly comparing starting, stopping, and continued use of OC, this prospective study shows that markers of ovarian reserve respond differently to changes in OC use status. The present findings allow a more nuanced interpretation of how biochemical and sonographic parameters reflect hormonally induced ovarian suppression and subsequent recovery.

Overall, the findings highlight that ovarian reserve markers, particularly AMH, should be interpreted with caution in the context of current or recent OC use, as they may reflect reversible functional changes rather than true underlying ovarian reserve. Among OC starters, serum AMH declined within three to six months, whereas AFC and OV did not. This suggests that AMH may be particularly sensitive to early ovarian suppression following the onset of OC. Previous studies investigating the impact of starting OC use on ovarian reserve markers have reported heterogeneous findings. Short-term studies (two to three weeks) generally found no changes in AMH levels (Andersen et al., 2011; Streuli et al., 2008), whereas Arbo et al. (2007) observed a decline in AMH, although starting OC occurred atypically during the luteal phase. Results from longer follow-up periods are similarly inconsistent. While AMH levels have been reported to remain stable (Li et al., 2011), reductions in AFC and OV despite unchanged AMH have also been observed (Somunkiran et al., 2007). In contrast, Kallio et al. (2013) reported a decline in AMH over nine weeks of continuous OC use, independent of administration route. These discrepancies likely reflect methodological heterogeneity, including differences in OC formulation, OC regimen (with or without HFI), timing of starting OC, follow-up duration, and AMH assays. The present study covers a mid-term observation window of three to six months. The stability of AFC and OV may suggest that structural ovarian parameters are less responsive to suppression following the start of OC use within this time frame, although the relatively small sample size may have limited statistical power to detect subtle changes. Previous cross-sectional studies reported an overall lower AFC in OC users, but also observed a shift toward smaller antral follicle subclasses, with larger follicles being more strongly reduced, whereas smaller follicles appeared unchanged or even increased (Birch Petersen et al., 2015; Deb et al., 2012). Based on these findings, it may be speculated that early after starting OC use, ovarian suppression initially affects follicular development and size distribution before resulting in a measurable decline in total AFC. This may help explain the absence of significant AFC changes in starters during the follow-up period of the present study. Overall, these findings raise the possibility that biochemical changes reflected by AMH may become detectable earlier than ultrasound-based markers following the start of OC use.

In contrast, stopping OC use was followed by a pronounced increase in all ovarian reserve markers within three to six months. AMH, AFC and OV increased compared with baseline values at S1 and relative to long-term OC users, indicating a quick recovery after cessation of hormonal suppression. These findings are consistent with earlier studies reporting normalization of ovarian reserve markers within a few menstrual cycles after stopping OC use (Landersoe, Birch Petersen, et al., 2020; Letourneau et al., 2017; van den Berg et al., 2010). The observed recovery is biologically plausible. AMH is produced by granulosa cells of preantral and small antral follicles (Weenen, 2004), reflecting the activity of this follicular pool. After stopping OC intake, removal of hypothalamic-pituitary-ovarian axis suppression allows renewed gonadotropin stimulation, promoting follicular growth and increased AMH production. The accompanying increase in AFC supports this interpretation, as progression of follicles into the antral size range (2–10 mm) becomes measurable. Given that follicular development from preantral stages to small antral follicles requires approximately 70 days (Gougeon, 2010), the timing of reassessment in the present study after at least three spontaneous menstrual cycles (corresponding to three to six months), is well aligned with this time frame. The increase in OV further reflects this renewed follicular activity, as ovarian size typically increases with a growing number of antral follicles.

While the present study design does not allow a detailed characterization of the dynamics after stopping OC using repeated measurements, as previously performed in longitudinal studies (Landersoe, Birch Petersen, et al., 2020; van den Berg et al., 2010), the results nevertheless provide robust evidence that recovery of ovarian reserve markers occurs within the first few spontaneous cycles. Cross-sectional data further suggest that previous OC use, even when long-term, is not associated with persistently reduced ovarian reserve. No differences in AMH levels approximately one year after stopping OC use compared with never-users have been reported (Kucera et al., 2016), and no association between prior OC use and lower AMH concentrations has been found, independent of duration of use (Bernardi et al., 2021). Together, these findings support the reversibility of OC-related suppression of ovarian reserve markers.

Among long-term OC users, markers of ovarian reserve remained stable over the study period. AMH, AFC and mean OV remained largely unchanged from baseline to follow-up, suggesting that OC-related effects on ovarian reserve markers were already established prior to study entry and did not further evolve during the relatively short observation period. Research regarding a duration-dependent decline during continued OC use is mixed. While some studies reported weaker ovarian reserve parameters with longer duration of use (Bentzen et al., 2012; Langton et al., 2022), others found no significant association after adjustment for age (Birch Petersen et al., 2015; Johnson et al., 2014) or observed such differences only after long-term use exceeding 20 years (Dólleman et al., 2013). Taken together, these findings suggest that suppression of ovarian reserve markers during continued long-term OC use may not be progressive but instead reaches a relatively stable suppressed state. This could explain why no further decline was observed during the follow-up in the present study. Accordingly, a pronounced decline in AMH, as reported during the early phase after starting OC use, would not be expected in this group. Although small decreases in AFC and OV were observed, these changes did not provide clear evidence of longitudinal change and are likely attributable to physiological intercycle variability (Ulrich & Marsh, 2019) or minor interobserver-related measurement variability. To our knowledge, no previous longitudinal study has specifically examined within-group changes among long-term OC users continuing OC use. Population-based analyses have reported lower AMH levels in current OC users compared with never-users, without evidence of an accelerated age-related decline (Werner et al., 2025). However, such studies cannot address intraindividual changes under continued long-term OC use. In this context, the present longitudinal data provide complementary evidence for the stability of ovarian reserve markers during ongoing OC exposure.

Overall, the findings of the present study have important implications for fertility counselling and the clinical interpretation of ovarian reserve testing. AMH appears particularly sensitive to both starting and stopping OC, whereas AFC and OV primarily reflect recovery after stopping. Consequently, assessment of ovarian reserve during active OC use or shortly after stopping OC carries a risk of misclassification. Transient suppression of AMH or AFC under hormonal contraception may therefore lead to an overestimation of diminished ovarian reserve, especially when diagnostic thresholds such as those proposed in the POSEIDON criteria are applied (Alviggi et al., 2016). It should be emphasized that AMH and AFC primarily function as prognostic markers of ovarian response within ART procedures rather than as direct measures of a woman’s natural fertility potential. Although AMH is associated with cumulative live birth rates after IVF or ICSI, no thresholds exist that reliably exclude the possibility of live birth (Peigné et al., 2023). Concerning fecundability, AMH is a poor predictor (Hagen et al., 2012), whereas age remains the strongest determinant of reproductive potential (Cedars, 2022). In line with these limitations, AMH has also been discussed as a marker of ovarian aging and timing of menopause. Although lower age-adjusted AMH values are associated with earlier menopause, individual prediction based on single measurements, particularly under hormonal contraception, remains highly uncertain (Nelson, Davis, et al., 2023). AMH should therefore not be used as a standalone tool in clinical practice to predict a woman’s reproductive lifespan.

Assessment of ovarian reserve may nevertheless be considered in selected women prior to starting OC use, as hormonal contraception may mask a pre-existing diminished ovarian reserve or early primary ovarian insufficiency (Kushnir et al., 2014). At the same time, there is no evidence that long-term use of OC has a lasting negative impact on fertility in the general population. Both short- and long-term use have been associated only with a temporary delay to conception after stopping OC (Mikkelsen et al., 2013). This finding is highly relevant for contraceptive counselling and should be clearly communicated to women concerned about future fertility.

While the prospective design, standardized timing of assessments, and standardized laboratory and ultrasound procedures strengthen the methodological robustness of this study, several limitations should be considered. First, subgroup sample sizes were relatively small, particularly among OC starters, which may have limited statistical power to detect more subtle changes, especially in sonographic parameters. Second, the restriction to a young and otherwise healthy population may limit the generalizability of the findings to older women or more diverse clinical settings. Third, ovarian reserve markers were assessed at only one follow-up time point, precluding detailed analysis of the dynamics after starting or stopping OC use. Future studies with larger samples, and repeated follow-up assessments over longer time periods are needed to better define the trajectory and clinical relevance of these changes.

In summary, starting OC use is associated with an early decline in AMH, whereas stopping OC intake leads to rapid recovery of AMH, AFC, and OV within a few menstrual cycles. Ovarian reserve markers remain largely stable during continued long-term OC use. These findings highlight the dynamic and reversible nature of OR markers under OC and emphasize the importance of accounting for current or recent OC use when interpreting these parameters in both clinical practice and research.

## Acknowledgement

We thank Sandra Beinbauer, Judith Scharpf, Lisa Stark and Grete Moeller for their help with data acquisition. The study, UB and MK were supported by the International Research Training Group: Women’s Mental Health Across the Reproductive Years (IRTG 2804), funded by the DFG; grant number: GRK 2804/1.

## Author contributions

MH, NBK and MK were responsible for the study concept and design. UB and MK collected data under supervision by MH and NBK. UB and MH conceived the method of this paper and UB and MK processed the data. UB performed the data analysis and MK contributed to analyses. UB wrote the manuscript. All authors contributed to the interpretation of findings, provided critical revision of the manuscript for important intellectual content and approved the final version for publication.

## Data Availability

De-identified individual participant data that support the findings of this study may be made available upon reasonable request to the corresponding author, subject to a data use agreement and approval in accordance with ethical and institutional data protection requirements.

