## Supplementary Tables S1-S3 for "Effects of Starting and Stopping Combined Oral Contraceptives on Markers of Ovarian Reserve"

### Supplementary Material

**Table S1**

*Descriptive values for right and left ovarian volume before and after starting, stopping, or continuing OC use*

| | Starters | | Stoppers | | Long-term users | |
|---|---|---|---|---|---|---|
| | **S1** | **S2** | **S1** | **S2** | **S1** | **S2** |
| **Right OV (mL)** | | | | | | |
| *Md* | 4.7 | 4.2 | 2.8 | 5.5 | 3.5 | 1.9 |
| Min–Max | 1.4–10.1 | 2.5–15.3 | 0.8–5.3 | 2.8–10.0 | 0.8–6.5 | 1.1–11.2 |
| **Left OV (mL)** | | | | | | |
| *Md* | 5.3 | 2.7 | 3.2 | 5.4 | 2.6 | 2.1 |
| Min–Max | 2.3–13.3 | 1.8–5.1 | 0.8–6.6 | 1.1–6.6 | 0.8–6.6 | 0.8–5.9 |

*Note.* Two ovarian volume measurements at S2 were excluded because of sonographically detected ovarian cysts.

**Table S2**

*Number of available AMH, AFC, and OV measurements by assessment and group*

| Variable | Group | S1 (n) | S2 (n) |
|---|---|---|---|
| **AMH** | Starters | 12 | 9 |
| | Stoppers | 16 | 10 |
| | Long-term users | 26 | 26 |
| **AFC** | Starters | 10 | 9 |
| | Stoppers | 14 | 8 |
| | Long-term users | 26 | 26 |
| **Mean OV** | Starters | 10 | 8 |
| | Stoppers | 15 | 8 |
| | Long-term users | 26 | 26 |

*Note.* Values reflect the number of observations included in the respective statistical analyses after accounting for missing data and exclusions of ovarian volume measurements due to sonographically detected ovarian cysts.

**Table S3**

*Linear mixed models for the analysis of changes in AMH, AFC, and OV between S1 and S2*

| | Predictor | *b* | *SE* | 95% CI | *df* | *t* | *p* |
|---|---|---|---|---|---|---|---|
| **AMH** | (Intercept) | 3.25 | 0.41 | [2.44, 4.04] | 59.35 | 7.98 | <.001*** |
| | S2 (Long-term users) | -0.28 | 0.27 | [-0.84, 0.28] | 41.25 | -0.98 | .333 |
| | S2 × Starters | -0.97 | 0.56 | [-2.06, 0.12] | 42.84 | -1.73 | .091• |
| | S2 × Stoppers | 2.85 | 0.54 | [1.81, 3.90] | 43.65 | 5.34 | <.001*** |
| **AFC** | (Intercept) | 10.12 | 0.67 | [8.82, 11.43] | 74.29 | 15.17 | <.001*** |
| | S2 (Long-term users) | -1.50 | 0.75 | [-2.98, -0.02] | 37.90 | -2.00 | .054• |
| | S2 × Starters | -0.21 | 1.49 | [-3.14, 2.71] | 42.46 | -0.14 | .887 |
| | S2 × Stoppers | 5.38 | 1.47 | [2.49, 8.27] | 44.76 | 3.65 | <.001*** |
| **Mean OV** | (Intercept) | 1.02 | 0.10 | [0.82, 1.22] | 70.57 | 9.85 | <.001*** |
| | S2 (Long-term users) | -0.19 | 0.10 | [-0.40, 0.02] | 39.56 | -1.77 | .084• |
| | S2 × Starters | -0.01 | 0.22 | [-0.44, 0.42] | 45.50 | -0.05 | .964 |
| | S2 × Stoppers | 0.85 | 0.21 | [0.44, 1.26] | 46.09 | 4.09 | <.001*** |

*Note.* Long-term users served as the reference group. The main effect “S2 (Long-term users)” describes the change in AMH, AFC, and OV values from S1 to S2 in the reference group. Interaction terms indicate the group-specific deviation from this change for starters and stoppers, respectively. The models controlled for centered age (years) and centered BMI (kg/m²). The mean OV was log-transformed (raw values in mL).

•$p < .10$, *$p < .05$, **$p < .01$, ***$p < .001$.
